# Risk Prediction In Long Term Kidney Transplant Recipients – Model Development Using Apelinergic Markers And Machine Learning Tools

**DOI:** 10.1101/2024.05.29.24308114

**Authors:** Krzysztof Batko, Anna Sączek, Małgorzata Banaszkiewicz, Jolanta Małyszko, Ewa Koc-Żórawska, Marcin Żórawski, Karolina Niezabitowska, Katarzyna Siek, Alina Bętkowska-Prokop, Marcin Krzanowski, Katarzyna Krzanowska

## Abstract

**Introduction:** Limited tools exist for predicting kidney function in long-term kidney transplant recipients (KTRs). Elabela and apelin are APJ receptor agonists that constitute the apelinergic axis, which is a recently discovered system regulating vascular and cardiac tissue, in opposition to renin-angiotensin-aldosterone.

**Methods:** Longitudinal, observational cohort of 102 KTRs who maintained graft function ≥24 months, with no acute rejection history or current active or chronic infection. Serum apelin, elabela, fibroblast growth factor 23 (FGF-23) and α-Klotho were tested using enzyme-linked immunoassay and compared with a control group of 32 healthy volunteers.

**Results:** Median (IQR) follow-up time was 83 (42, 85) months. Higher serum FGF-23 and elabela, but lower α Klotho concentrations were observed in KTRs. Most KTRs had stable trajectories of renal function. All candidate markers were significantly associated with mean two-year eGFR over follow-up, which itself was validated respective to ‘death with functioning graft’ censored dialysis requirement. Using a cross-validation approach, we demonstrated eGFR at initial visit as the most salient predictor of future renal function. Machine learning models incorporating both clinical and biochemical (candidate markers) assessments were estimated to explain 15% of variance in future eGFR when considering eGFR-independent predictions.

**Conclusions:** Utilization of machine learning tools that incorporate clinical information and biochemical assessments, including serum amrkers of the apelinergic axis, may help stratify risk and aid decision making in the care of long term KTRs.

## 1. Introduction

Chronic kidney disease (CKD) is an irreversible disorder with a slowly progressive course that is tied to excess morbidity. Studies estimate its global prevalence at over 10% within the general population, with non-linear growth and even higher rates among the elderly^1^. Cardiovascular (CV) disease remains one of the leading causes of death across the whole spectrum of CKD. CV-related mortality is starkly elevated in end-stage CKD, especially in subjects requiring renal replacement therapy (RRT), but also remains significantly elevated after kidney transplant (KTx)^2^.

Apelin (APLN) and elabela (ELA; toddler or apela) are two endogenous ligands of the APJ, a G protein coupled receptor tied to several intracellular transduction pathways affecting adenylyl cyclase activity, ion gradients (including calcium shift) and nitric oxide synthesis^3,4^. APJ shows widespread expression in multiple tissues, including cardiac, vascular and renal organs^1,5,6^. The APJ receptor shows modest homology and comparable tissue distribution to angiotensin receptor type 1, which has led to speculation of physiological role as a counterbalance to the renin-angiotensin-aldosteron (RAAS) axis, alterations of which result in accelerated atherosclerosis and various organ disorders^7–10^.

APLN and ELA are 77- and 54-amino acid preproproteins cleaved by tissue proteases into shorter peptides, which are secretable^11^. While the C-terminal APLN sequence determines receptor binding, the N-terminal interacts with the APJ receptor, which may explain variability in tissue affinity of specific isoforms^1,12^. Most immunological assays target the C-terminal fragment of ELA, which precludes differentiation of specific isoforms. Immunoassay remains the only robust method of testing ELA concentrations^13,14^. Contrary to prior hypotheses suspecting an ELA-specific receptor, ELA has been demonstrated to act as a ligand of the APJ receptor, with near selective expression within the vascular endothelium of the kidney^15–17^.

In contrast to the early post-Ktx period, long-term kidney transplant recipients (KTRs) that maintain stable graft function represent a distinct population that is susceptible to CV and metabolic diseases, which represents the major risk factor for graft loss and/or death^18^. Developing prognostic tools for these patients is of particular interest due to the paucity of measures that are validated specifically in such populations. Due to its strong link with CV disease, the recently discovered apelinergic axis represents an important regulator of tissue homeostasis and potential source of biomarkers.

This study was undertaken to develop a predictive model for future renal function in stable, long-term KTRs. This analysis includes an exploratory investigation of the potential relevance of apelinergic markers in serum, including their association with clinical co-variates and definitive post-KTx outcomes. While models derived based on large population of KTRs have been previously published, the population of stable, long-term KTRs represent a unique group, for which the validity and relevance of published risk scores is uncertain.

## 2. Methods

### 2.1 Patients

This was a longitudinal, observational cohort study, which enrolled 102 consecutive patients under ambulatory care at the University Hospital in Kraków, Poland. The recruitment process had an annual timeframe between September 1, 2016 and October 31, 2017, which was pre-defined to last at least one year due to slow rate of recruitment. Subjects were considered for study participation only if they were long-term KTRs, which was defined as having maintained allograft function for at least 24 months. Episodes of acute rejection remain one of the major risk factors for chronic rejection^19^, therefore patients in whom history of acute rejection was recorded (whether cellular, humoral or vascular), were not included. We further excluded patients with infection, both active (ie signs or symptoms of any acute infection at presentation) or chronic (based on serological evidence in routine work-up, eg viral hepatitis, HIV). Due to altered mineral-bone imbalance, individuals with prior parathyroidectomy status or a history of malignancy (excluding non-melanoma skin cancer), were also excluded due to the presence of (potential) salient interfering factors. The recruitment process is illustrated by a flowchart (Figure S1). The inclusion and exclusion criteria are consistent with our earlier studies.

### 2.2 Follow-up

Over time, electronic medical care records were screened manually by four physicians (K.B., M.B., A.S., K.N.) to identify visits at relatively equally spaced intervals, with a pre-defined range of 3 to 6 months apart. The total follow-up time for renal function measures was comparable for most patients, with a median (IQR) timeframe of 23.6 (22.8, 24.4) months and range of 21.7-25.3 months. The occurrence of events of poor prognosis, such as allograft loss (defined as permanent dialysis transfer or re-transplantation listing) or death was determined through in person or telephone contact with the patient, family and/or dialysis center. This process was carried out successively until December 30, 2023 (censoring date).

### 2.3 Covariates

Clinical data, which included age, sex, body mass index (BMI), KTx characteristics (primary etiology of kidney disease, immunosuppresive treatment, history of delayed graft function) and co-morbidity (CV disease, hypertension, diabetes and dyslipidemia) were gathered. CV disease was defined as records of either atherosclerotic CV disease (ie, a composite of coronary artery disease (CAD), prior myocardial infarction (MI) or prior stroke) or heart failure (HF).

Nearly all patients were treated with triple immunosuppressive therapy (glucocorticoids, mycophenolate mofetil and a calcineurin inhibitor) and none had history of induction pre-treatment. Glomerulonephritis, autosomal dominant polycystic kidney disease and reflux nephritis were the most common primary causes of KTx. Information regarding death or dialysis/re-transplantation was determined based on in-person and/or telephone contact with the patient, family and/or dialysis center.

### 2.4 Biochemical testing

Peripheral venous cannulation was performed at presentation on the morning following an overnight fast into EDTA tubes, which were subsequently frozen at -70 C and stored until analysis. Standard biochemistry assays were performed on either of three automatic analyzers: Hitachi 917 (Hitachi, Japan); Modular P (Roche Diagnostics, Germany); Sysmex XE 2100 (Sysmex, Japan). For non-routine biochemical assays, testing was performed in batch, using commercially available immunoenzymatic kits according to manufacturer instructions. A healthy control group of 32 volunteers (16 female; 16 male), aged between 29 and 74 years (mean 50 years) was recruited using convenience sampling among willing physicians and medical personnel. A more detailed description of this control group is available elsewhere^20^.

Serum apelin was measured using Human Apelin ELISA Kit (EIAab Science, Wuhan, China), with a detectable level of 62.5 - 4000 pg/mL, sensitivity of 30 pg/mL, and intra- and inter-assay precision of <7% and <9%, respectively.

Serum elabela was measured using Cat. No. S-1508 ELABELA (Peninsula Laboratories, San Carlos, CA, USA), with a detectable level of 0-100 ng/mL, and intra- and inter-assay precision ∼10% and ∼15% (as reported^21^), respectively. No cross-reactivity with apelin has been reported.

Serum klotho was measured using Human Soluble α Klotho kit (IBL, Gunma, Japan), with a detectable level of 93.75 – 6 000 pg/mL, sensitivity of 6.15 pg/mL, and intra- and inter-assay precision of ∼3% and ∼7%, respectively.

Serum FGF-23 was measured using Human FGF-23 Intact ELISA kit (Immunotopics, San Clemente, USA), with a detectable level of 31.25-2000 pg/mL, sensitivity of 15 pg/ml, and intra- and inter-assay precision of ∼7% and ∼11%, respectively.

### 2.5 Bioethics statement

The conduct of this study was in adherence with the Declaration of Helsinki principles, as well as International Conference on Harmonization/Good Clinical Practice regulations. Study approval was obtained from the Bioethics Committee of the Jagiellonian University and all patients provided written informed consent prior to study inclusion.

### 2.6 Statistical analysis

Analyses were performed in R 4.3.2 (R Core Team, 2024, R Foundation for Statistical Computing, Vienna, Austria).

Nominal variables were summarized as counts and proportions (N, %). Variable distribution was assessed using density plots, the Shapiro Wilk test and skewness measures. Continuous variables were summarized as mean (standard deviation) or median (interquartile range), as deemed appropriate. Comparison across groups is performed using Welch’s t test (supplemented with permutation based testing) for continuous variables. The measure of association for two dichotomous variables was assessed with x^2^ test or Fisher’s exact test.

The nephro package was used to calculate estimated glomerular filtration (eGFR) according to the Chronic Kidney Disease-Epidemiology Collaboration (CKD-EPI) formula based on serum creatinine^22^. While the median may be conceptually more robust than the mean, due to very high correlation between both measures, the mean was selected.

Prognostic models were constructed using random forest, as a high performance machine learning model, using the *ranger* package within the *tidymodels* package. Due to generally low rates of missing data per variable (<10%), we utilized imputation using bagging techniques. Tuning was performed with grid search for hyperparameter selection using five times repeated tenfold cross-validation. Due to sample size considerations, feature selection was determined using an initial model with all potential covariates, further trimmed according to variable importance and expert knowledge. Permutation-based model breakdown techniques using the *vip* and *DALEX* package were utilized to analyze feature contribution to model prediction from a global and local perspective, respectively.

Statistical tests were two-tailed and P value < 0.05 was deemed statistically significant.

## 3. Results

### 3.1 Baseline clinical characteristics of kidney transplant recipients

This was a cohort of middle-aged adults, with mean (SD) age of 50.8 (14.5) years, and male predominance (N=72, 70.6%). Most patients were overweight, with mean (SD) body mass of 26.1 (4.68) kg/m^2^. Tacrolimus-based triple therapy regimens were most commonly utilized (N=67, 66.3%), followed by cyclosporine-based (N=22, 21.6%). History of delayed graft function, defined as dialysis requirement within 7 days in the early Tx period, was documented for every fifth patient (N=23, 22.6%). For mortality and kidney allograft loss, the median (IQR) follow-up time was 82.6 (42.1, 85.2) months. Most patients retained adequate kidney function at baseline, with mean eGFR upon presentation at 58.4 (22.6) ml/1.73m^2^/min, respectively.

Most subjects were hypertensive (N=92, 90.2%), with dyslipidemia and diabetes reported for 15 (14.7%) and 25 (24.5%) individuals, respectively. Nearly all diabetics (N=29, 28.4%) were characterized with post-transplant diabetes (N=24, 23.5%). Manifest CV disease was observed in every fourth patient (N=25, 24.5%). A comparison of clinical features according to allograft status at last available follow-up is shown in Table 1.

**Table 1.** Baseline demographic, clinical and biochemical characteristics compared among kidney allograft survivors and patients requiring dialysis transfer.

|  | Allograft survival (N=77) | Allograft dysfunction (N=25) | *P* value |
| --- | --- | --- | --- |
| Age, years | 52.03 (14.33) | 47.12 (14.71) | 0.14 |
| Male sex, N (%) | 52 (67.5%) | 19 (79.2%) | 0.24 |
| BMI, kg/m<sup>2</sup> | 25.81 (4.73) | 26.91 (4.50) | 0.32 |
| Baseline eGFR, ml/min | 64.38 (20.82) | 40.09 (17.94) | < 0.001 |
| CKD stage, N (%) |  |  | <0.001 |
| IV | 2 (2.6%) | 7 (28.0%) |  |
| IIIB | 14 (18.2%) | 13 (52.0%) |  |
| IIIA | 19 (24.7%) | 1 (4.0%) |  |
| II | 33 (42.9%) | 3 (12.0%) |  |
| I | 9 (11.7%) | 1 (4.0%) |  |
| CVD, N (%) | 16 (20.8%) | 9 (36.0%) | 0.12 |
| Hypertension, N (%) | 69 (89.6%) | 23 (92.0%) | 0.73 |
| MAP, mm Hg | 99.98 (11.93) | 103.03 (13.86) | 0.30 |
| Dyslipidemia | 10 (13.0%) | 5 (20.8%) | 0.35 |
| Diabetes, N (%) | 9 (11.7%) | 6 (24.0%) | 0.13 |
| Fasting glucose, mmol/l | 5.63 (1.39) | 5.75 (0.90) | 0.68 |
| Hemoglobin, g/L | 13.75 (2.12) | 13.92 (1.46) | 0.71 |
| FGF-23, pg/mL | 1.26 (1.57) | 3.28 (4.32) | <0.001 |
| $\alpha$ Klotho, pg/mL | 838.50 (652.55) | 536.55 (231.45) | 0.03 |
| Elabela, ng/mL | 33.66 (13.14) | 30.00 (9.08) | 0.20 |
| Apelin, pg/mL | 1446.82 (1173.79) | 1672.30 (1136.82) | 0.40 |

Abbreviations: blood pressure, BP; body mass index, BMI; estimated glomerular filtration,
eGFR; manifest cardiovascular disease, CVD; fibroblast growth factor type 23, FGF-23; mean
arterial blood pressure, MAP

When comparing with healthy controls, we observed higher concentrations of serum FGF-23 (mean difference [SE] 1055.39 (488.63), *P*<0.001) and elabela (mean difference [SE] 723.28 (252.36), *P*=0.003) in KTRs, alongside singificantly lower α Klotho (mean difference [SE]: -543.20 (146.59), *P*=0.02) levels, but apelin (mean difference [SE]: -138.26 (295.13), *P*=0.77) (see Figure 1).

**Figure 1.**
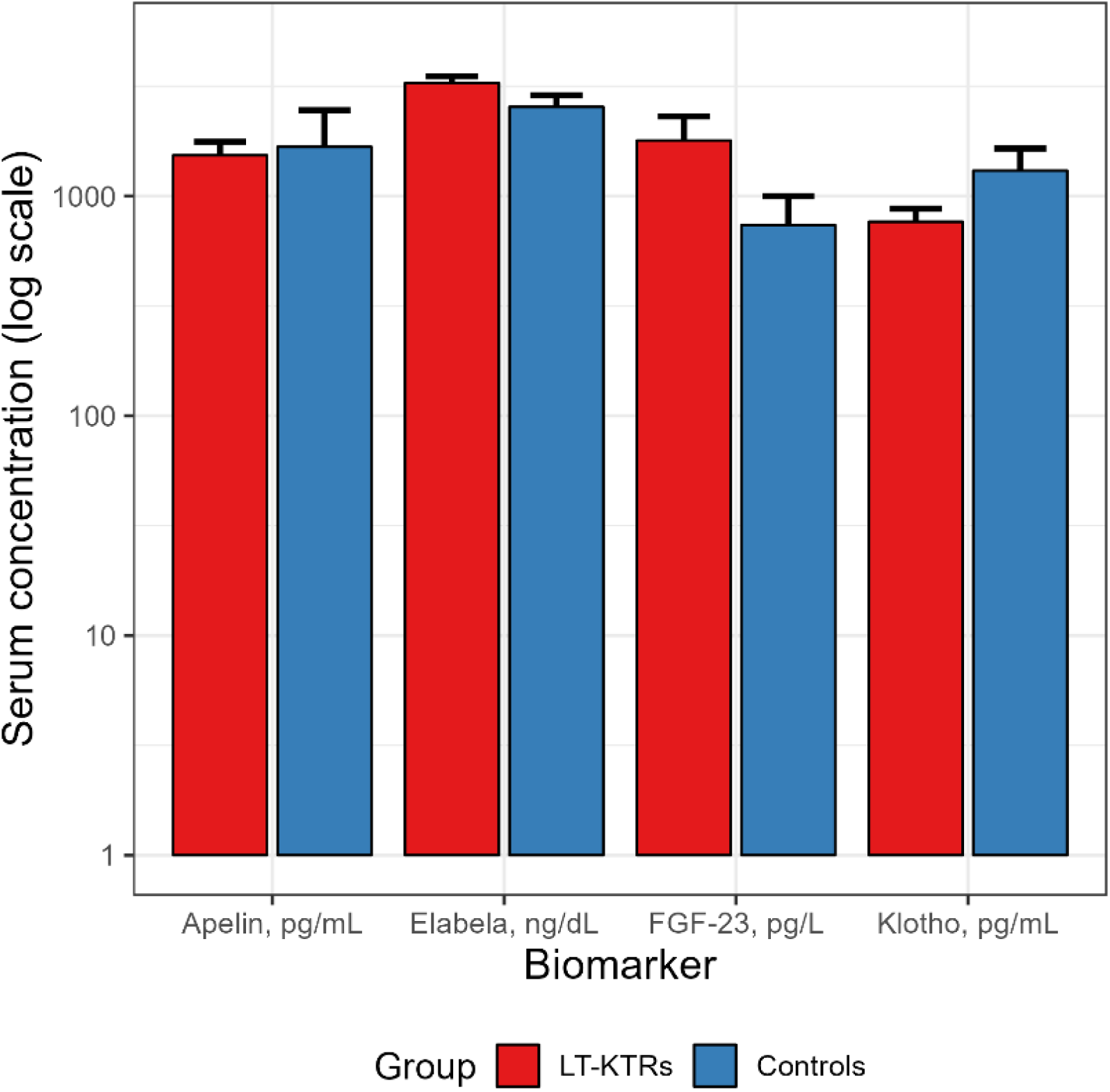
Differences in mean serum concentrations of apelin, elabela, fibroblast growth factor typ 23 and alpha klotho compared between kidney transplant recipients and healthy controls.

### 3.2 Relationships between kidney function, vascular and metabolic parameters and circulating concentrations of apelinergic peptides and mineral-bone hormones

This cohort included patients from a broad range of graft function at inception. When examining temporal changes in eGFR, we observed that patients who maintain allograft function have stable trajectories, as compared with subjects who (at a later date) required dialysis therapy and exhibited a gradual trend of eGFR decline (Figure 2, Panel A). In an age-adjusted logistic regression model (Somer’s Dxy 0.76, optimism corrected R^2^=0.52), higher mean eGFR over two-year follow-up was associated with significantly lower odds of graft loss (OR 0.04, 95% CI 0.01-0.15; *P*<0.001). The predicted probabilities are illustrated and adjusted for age, according to the interquartile cut-offs (Figure 2, Panel B).

**Figure 2.**
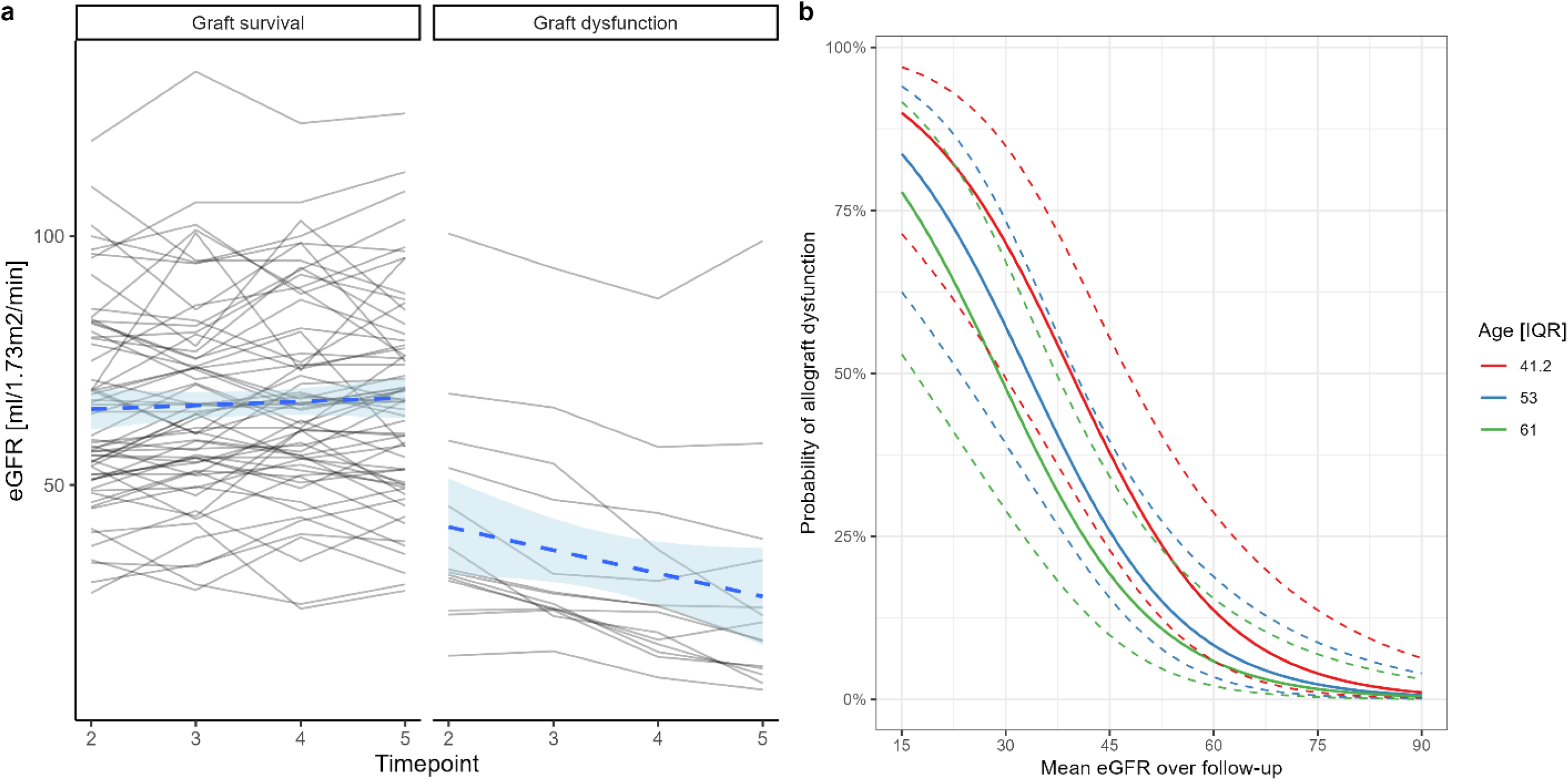
Temporal changes in renal function based on repeat eGFR testing and stratified by dialysis requirement (Panel A). Predicted probabilities of renal allograft loss based on mean eGFR values calculated over time and based on two different approaches (Logistic regression – Panel B; Cox PH – Panel C).

Baseline renal function showed no significant association with apelinergic markers (R=-0.10, *P* =0.36 for apelin, r=0.15, *P*=0.13 for elabela), in contrast to mineral bone peptides (R=-0.25, *P*=0.02 for FGF-23; r=0.34, *P*<0.001 for α Klotho, respectively). Only a weak and inverse linear trend could be observed between serum apelin and elabela (r=-0.18, *P*=0.08), as well as between FGF-23 and α Klotho (r=-0.15, *P*=0.14).

We observed significant relationships between circulating concentrations of apelinergic peptides and mineral-bone hormones with future eGFR over follow-up (Figure 3).

**Figure 3.**
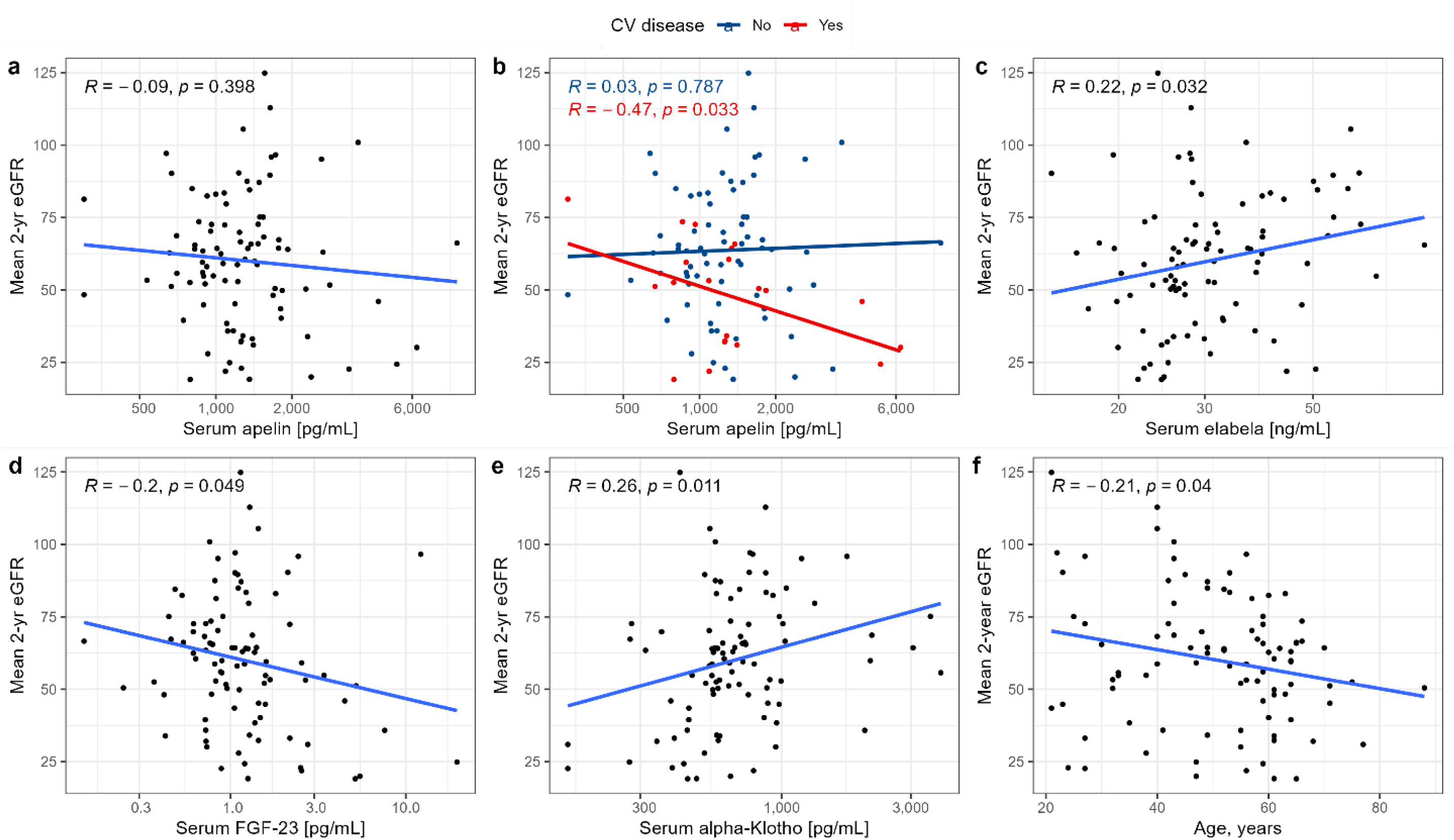
Relationship between mean eGFR over follow-up and baseline concentrations of the studied markers.

### 3.3 Developing a robust prognostic model for future kidney function

To understand the importance of baseline eGFR, a univariable random forest regression model was tuned and estimated to explain 87% of variance (R^2^ [SE]: 0.865 [0.01]) in future 2-year eGFR. This suggests the overarching importance of eGFR assessments over follow-up in stable KTRs. Therefore, mean eGFR over follow-up – baseline eGFR was adopted as a measure of kidney outcome agonistic of eGFR. This approach enables easier interpretation of other, clinically-relevant covariates.

For the final random forest model (agnostic of baseline eGFR), we observed stable performance with ability to explain 15% of variance (R^2^ [SE]: 0.146 (0.02) in future 2-year eGFR minus baseline eGFR). We also assessed the performance of this model based on aggreagated estimates over resamples, with a mean absolute error of 7.41 (0.194) and a root mean squared error of 9.01 (0.240). Global variable importance is presented in Figure 4 (left-hand panel), while prediction analysis for a sample clinical case illustrates how biochemical markers and clinical characteristics contribute to eGFR prognosis (right-hand panel).

**Figure 4.**
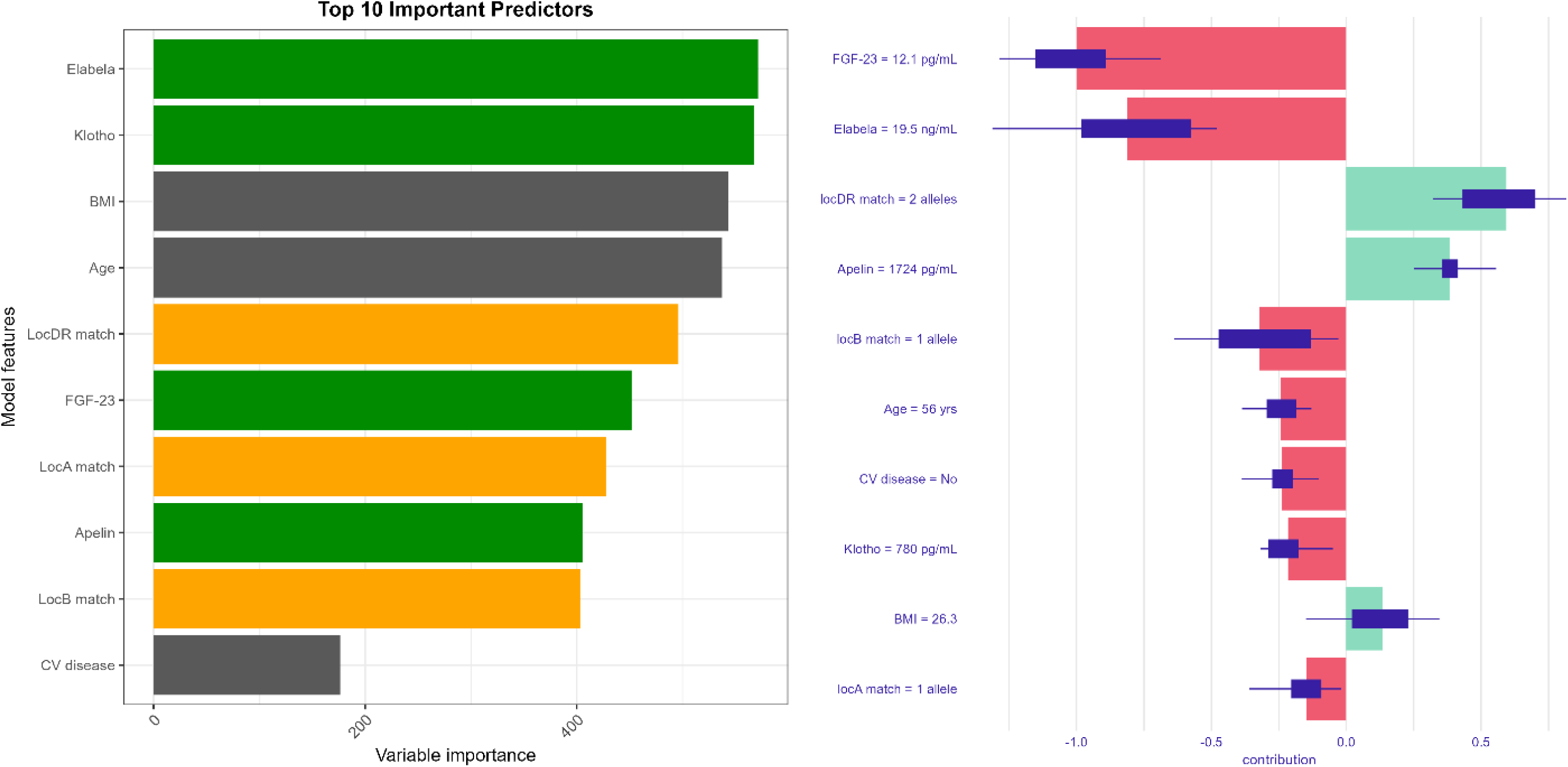
Global feature importance and local prediction breakdown for a real-life patient case.

When examining marginal effects of age and body-mass (Figure 5, Panel E & F), we observed a log-like relationship for age and non-monotonic contribution of BMI. It is probable that overall, older age and normal BMI are likely to exert a protective effect on future renal function. For APJ agonists, circulating apelin and elabela deficiency may be associated with worse prognosis for eGFR change, which contrasts with the effect of mineral-bone peptides.

**Figure 5.**
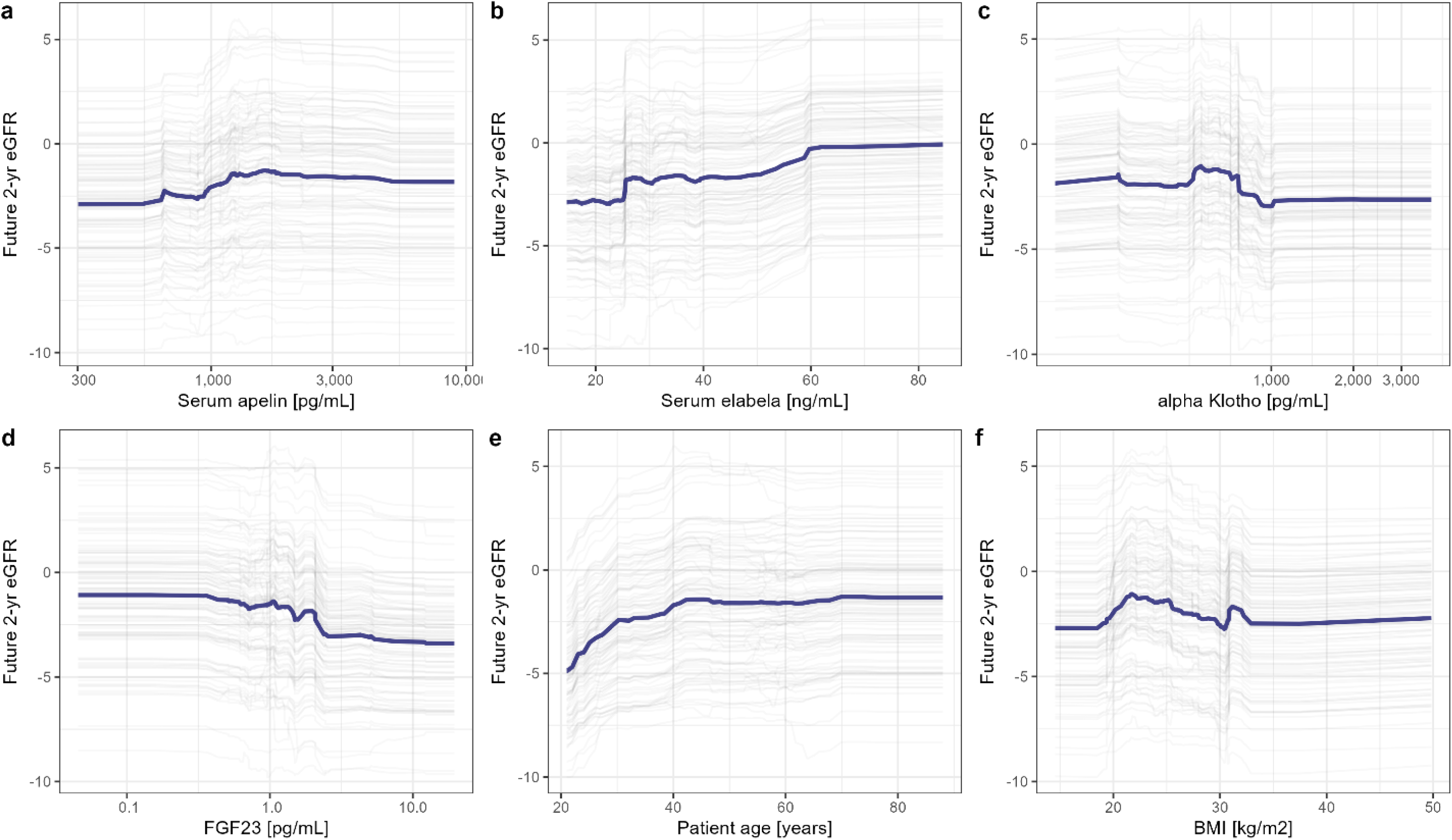
Partial dependence plots illustrating marginal effects of selected clinical features on model predictions.

## 3. Discussion

This was a longitudinal cohort study of long-term, stable KTRs, in whom we observed stable trajectories of graft function over two year follow-up. While post-KTx renal function is a major determinant of graft and patient survival^23^ and acute rejection remains a critical offender in the early stages, less is known about risk stratification in long-term KTRs. Epidemiological trends suggest an improvement in prognosis over recent years, which has been attributed to modern treatment schemes^24^. However, the case of stable, long-term KTRs is particularly interesting, as under the assumption of satisfactory HLA matching and compliance with immunosuppressive treatment, a number of different risk factors (including CV and metabolic) for allograft loss emerge in importance^19^.

There are several theoretical considerations that justify investigation of the apelinergic system as a prognostic measure in KTRs. Elabela and apelin show partially overlapping distribution throughout the vasculature^15,25^. Their joint receptor activation promotes vasodilation^15,26^ and increases cardiac contractility^15,27^, suggesting importance in a compensatory response to developing vascular disease. Meanwhile, within the kidney, APJ expression is more prevalent within glomeruli, indicating its hypothetical utility as a measure of glomerular, rather than tubular injury^16,28,29^. Kidney injury results in apelin expression^30^, while elabela mRNA levels are markedly reduced in ischemia-reperfusion models^31^, suggesting an inverse role in the reparative response. Both apelinergic agonists are attributed cardio-^32,33^ and renoprotective^10,34,35^ roles, though the activity of elabela is describes as more potent^31^. Thus, alterations in elabela, rather than apelin, are expected to parallel impairment within the kidney vasculature.

Among the investigated markers, significantly higher concentrations of elabela, FGF-23 and lower α Klotho concentrations were observed among KTRs, as compared with healthy controls. Both elabela and α Klotho serum concentrations showed a significant relationship with future eGFR, while a similar association for apelin was noted only among individuals with overt CV disease. Using a robust machine-learning technique, we constructed predictive models to assist in understanding the prognostic contribution of these novel markers, which may be comparable to clinically-established covariates. However, the overarching importance of eGFR at prior visits is also evident for patients with long-term, stable kidney allograft.

In patients with kidney disease, plasma apelin concentrations were first described by Malyszko et al.^36^. Lower concentrations of apelin were tied to CVD, with ventricular dimensions in echocardiography as its major determinants, speculated to parallel to the extent hemodynamic overload^36^. However, these were prevalent dialysis subjects. Another study in autosomal dominant polycystic kidney disease noted an independent association between apelin and renal function decline, speculating it may parallel dehydration status and suggesting its utility as a prognostic measure, beyond the information derived from eGFR estimates^37^. In patients with type 2 diabetes, serum apelin concentrations were markedly higher (compared with healthy subjects) and associated with albuminuria, an early indicator of diabetic nephropathy. Experimental evidence suggests apelin may modulate the permeability and proliferation capacity of glomerular endothelial cells, which may affect hyperfiltration of glomeruli^38^. In contrast, a report by Lu et al. showed no significant differences in apelin concentrations across patients with different stages of CKD when comparing age- and sex-matched controls^39^. Further work is necessary to clarify the relevance of circulating apelin concentrations.

Prior studies have suggested that elabela may be associated with progression of nephropathy^39^. Data from two study groups suggests that aside from selective expression within the kidney, it is localized tubular, rather than glomerular compartments^16,40^. Through its interaction with the Gi signaling pathway, elabela is a likely regulator of fluid homeostasis^17^, though it may also act to antagonizes aldosterone-stimulation of kidney tissue^41^. Numerous experimental reports identify elabela expression as an effective countermeasure to renal injury^15,40^, though whether this is mechanistically linked to cell cycle and/or anti-inflammatory effects^6^ remains unclear.

Apelinergic activity can also be ascribed a protective role in arterial calcification due to its regulation of smooth muscle cell differentiation into osteoblast lines^42^ and attenuation of calcification processes^43^. While mineral-bone peptides (FGF-23 and α Klotho) did not show any apparent association with apelinergic markers, they contributed to model predictions and shared an association with renal function over follow-up. We hypothesize that FGF-23 excess and alpha Klotho deficiency slowly develops in post-Ktx patients, in a manner akin to progressing CKD. Therefore, its supplemental prognostic value in KTx recipients may parallel recognition as risk factor of kidney dysfunction in the general population^44^.

Traditional CV risk factors, with underlying metabolic disorders and excessive RAAS activation, are processes that promote allograft dysfunction^45^. Apelin shares a close relationship with TNF α expression in adipose tissue, which has lead to its consideration as a potential bridge mediator between inflammation and insulin resistance in obesity^7,46^, though body-mass is likely not a major determinant of its circulating forms^47^. In patients with type 2 diabetes, serum elabela levels are inversely tied to proteinuria and creatinine elevation^48^, while experimental evidence suggests a protective role of elabela that reduces podocyte apoptosis^49^. Antagonism between the RAAS and APJ axes has been previously described^50,51^ and a simplified schematic is provided for the reader in Supplementary Figure S2. Importantly, on a systemic level, apelinergic activity may only be marked under conditions of tissue stress^52,53^, as demonstrated by experimental studies of myocardial contractility in failing hearts^54,55^. This may explain, at least in part, the conflicting findings of patient-level studies.

In model construct we utilized novel, potential markers of recently discovered biologic processes that can shape cardiorenal disease. Interpretation of circulating concentrations and causal cross-attribution, respective to their purported biologic roles, remains tentative. The apelinergic system remains incompletely understood, particularly in the post-KTx setting. We assessed both apelin and elabela in human serum using enzyme-linked immunosorbent assays, as has been performed by others. The potential variability in bioactivity and tissue affinity of specific molecular isoforms, as well as different sources of circulating peptides requires clarification in future studies. Interactions within signaling pathways tied to vascular disease (e.g., shear stress, endothelial dysfunction) or immune dysregulation may further occlude our conceptual understanding of changes in serum/plasma concentrations of specific markers. Thus, the results of the present study may be influenced by interaction with unknown clinical characteristics or incomplete understanding of biologic activity.

Many studies are limited by utilization of binary outcomes derived from repeated eGFR measures. Inter- and intra-individual variation in eGFR assessments, as well as timepoint number and temporal relation should be considered. To improve clinical interpretation, we defined the response variables as mean sequential eGFR and last available eGFR. For both outcomes, a strong association with ‘death with functioning graft’ censored dialysis was verified. To limit bias of the model predicting last available eGFR, we also utilized internal validation based on resampling techniques.

## 4. Conclusions

Discovering new biomarkers with prognostic potential and developing a risk model for renal function loss in KTRs holds high clinical importance, as no single parameter is sufficient to identify patients at greatest risk for allograft loss, and routine biochemical and clinical tests are often insufficient to predict graft failure accurately. The developing promise for apelinergic therapeutics in cardiac and renal diseases suggests another, supplemental role for apelin and elabela, as candidate biomarkers of a multisystem axis that counteract the effects of RAAS^1,6,10^.

Following the early post-KTx stage, allograft loss is difficult to predict and complex models involving demographic, clinical and biochemical data are warranted. This was an observational cohort study of long-term, stable KTRs, which examined temporal trajectories of renal function based on sequential eGFR assessments. Candidate biomarkers tied to apelinergic activity and mineral-bone imbalance were evaluated, with theoretical justification derived from modulatory activity within vasculature and renal tissue. Circulating concentrations of serum elabela, apelin, FGF-23 and α Klotho concentrations were significantly associated with mean eGFR over two year follow-up. After accounting for kidney function at initial visit, a random forest model combining basic clinical information and biomarker levels was estimated to account for 14% of variance in future eGFR.

## Conflict of interest statement

The authors declare that there is no conflict of interest regarding the publication of this paper.

## Data Availability

All relevant data are within the manuscript and its Supporting Information files.

## Acknowledgments

This study was conducted with the use of equipment purchased by Medical University of Bialystok as part of the RPOWP 2007–2013 funding, Priority I, Axis 1.1, contract No. UDA-RPPD.01.01.00-20-001/15-00 dated 26 June 2015.

## Funding sources

This study was supported by a statutory grant from the Jagiellonian University Medical College (N41/DBS/000193; to K.K.). This study was performed using equipment of the Medical University of Bialystok acquired via the RPOWP 2007-2013 funding grant, Priority I, Axis 1.1, contract No. UDA-RPPD.01.01.00-20-001/15-00 dated to 26.06.2015.

## Author contributions

All authors contributed significantly to the present report. All authors performed critical revision and provided acceptance of this paper. Conceptualization, K.B., K.K.; Data curation, K.B., K.N., K.S., A.S., M.B., J.M., K.K., M.K., E.K.-Ż., M.Ż., A.B-P.; Formal analysis, K.B., K.N., K.S., A.S., M.B., J.M., K.K., M.K., E.K.-Ż., M.Ż., A.B-P.; Funding acquisition, K.K. Investigation, Methodology, K.B., K.N., K.S., A.S., M.B., J.M., K.K., M.K., E.K.-Ż., M.Ż., A.B-P.; Software, K.B.; Supervision, J.M., M.K., K.K.; Writing—original draft, review and editing, K.B., K.N., K.S., A.S., M.B., J.M., K.K., M.K., E.K.-Ż., M.Ż., A.B-P. All authors have read and agreed to the published version of the manuscript.

## Abbreviations

APJ: apelin/elabela receptor
BMI: Body mass index
CV: Cardiovascular
CKD: Chronic kidney disease
CKD-EPI: Chronic Kidney Disease-Epidemiology Collaboration
eGFR: Estimated glomerular filtration
HF: Heart failure
KTx: Kidney Transplant
KTRs: Kidney Transplant Recipients
MI: Myocardial infarction
RRT: Renal replacement therapy
RAAS: Renin angiotensin aldosteron

---

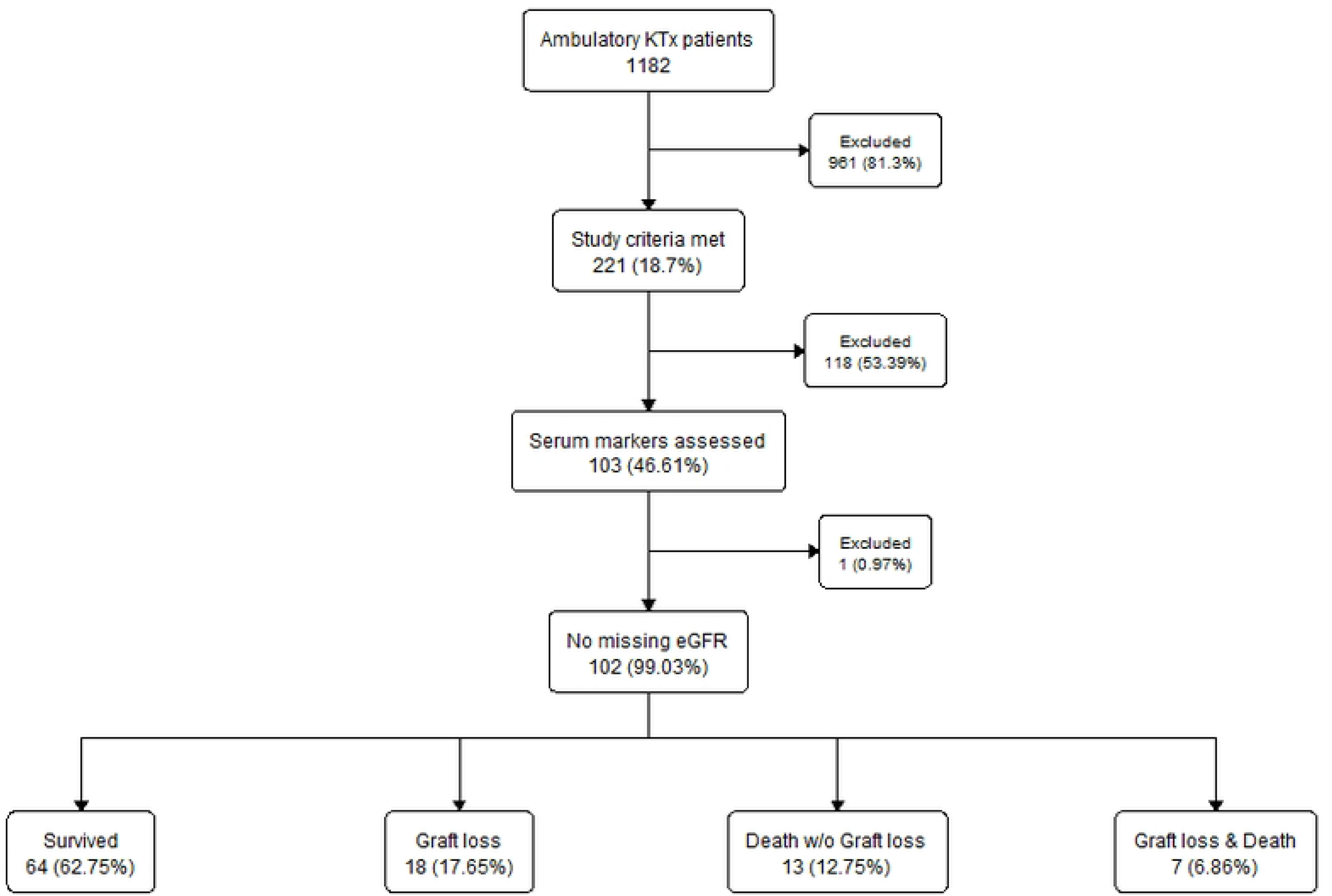

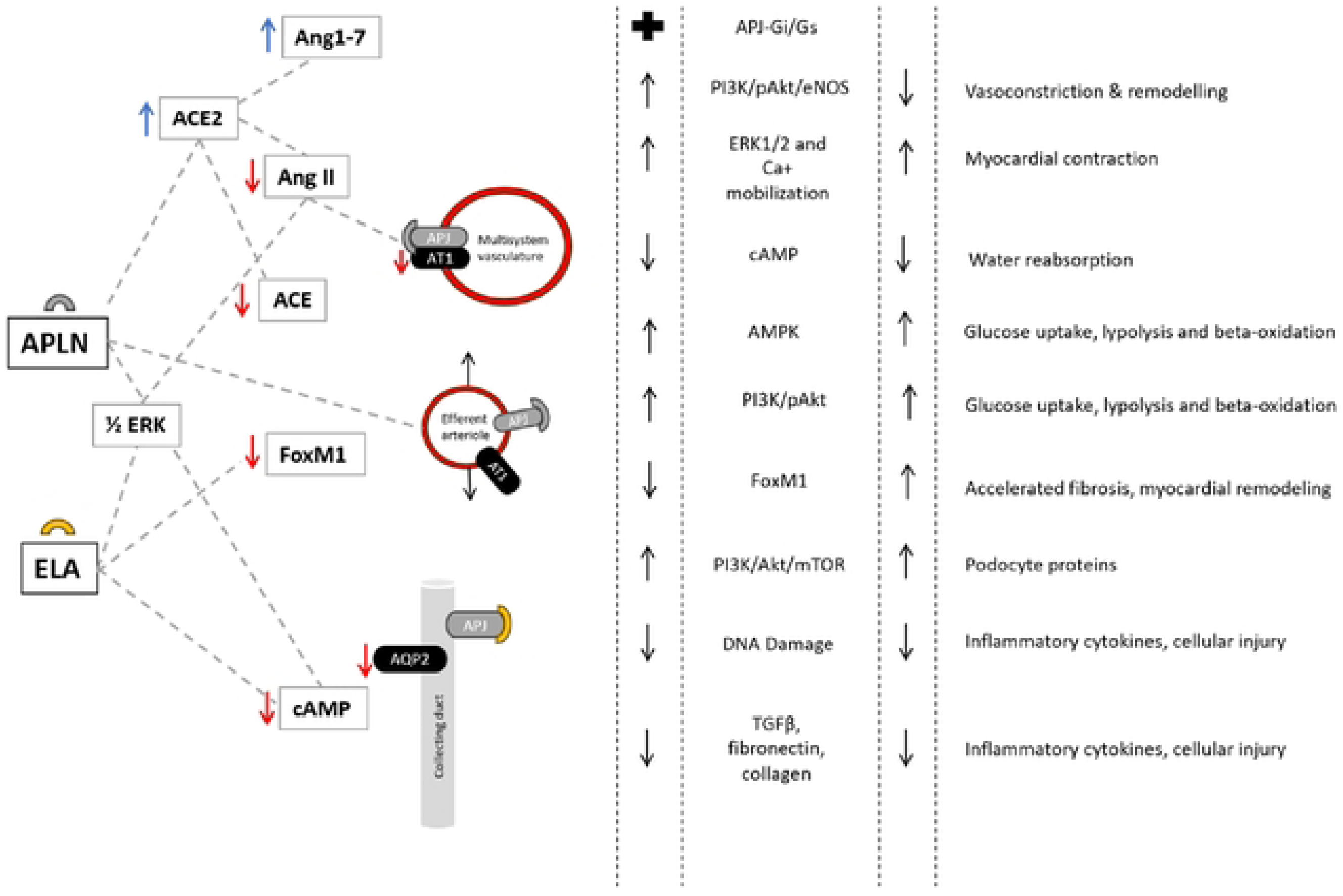

## Notes

### Competing Interest Statement

The authors have declared no competing interest.

### Funding Statement

Yes

## References

1. Chapman, F. A. et al. The therapeutic potential of apelin in kidney disease. Nat Rev Nephrol 17, 840– 853 (2021).

2. Kim, J. E. et al. De novo major cardiovascular events in kidney transplant recipients: a comparative matched cohort study. Nephrology Dialysis Transplantation gfac144 (2022) doi:10.1093/ndt/gfac144.

3. Szokodi, I. et al. Apelin, the novel endogenous ligand of the orphan receptor APJ, regulates cardiac contractility. Circ Res 91, 434–440 (2002).

4. Wang, B., et al. The Involvement of Chronic Kidney Disease and Acute Kidney Injury in Disease Severity and Mortality in Patients with COVID-19: A Meta-Analysis. Kidney Blood Press Res 46, 17–30 (2021).

5. Boucher, J. et al. Apelin, a newly identified adipokine up-regulated by insulin and obesity. Endocrinology 146, 1764–1771 (2005).

6. Zheng, Q. et al. The role of Elabela in kidney disease. Int Urol Nephrol 53, 1851–1857 (2021).

7. Wysocka, M. B., Pietraszek-Gremplewicz, K. & Nowak, D. The Role of Apelin in Cardiovascular Diseases, Obesity and Cancer. Front Physiol 9, 557 (2018).

8. Yu, X.-H. et al. Apelin and its receptor APJ in cardiovascular diseases. Clin Chim Acta 428, 1–8 (2014).

9. Huang, Z., Wu, L. & Chen, L. Apelin/APJ system: A novel potential therapy target for kidney disease. J Cell Physiol 233, 3892–3900 (2018).

10. Day, R. T., Cavaglieri, R. C. & Feliers, D. Apelin retards the progression of diabetic nephropathy. Am J Physiol Renal Physiol 304, F788–800 (2013).

11. Reichman-Fried, M. & Raz, E. Small proteins, big roles: the signaling protein Apela extends the complexity of developmental pathways in the early zebrafish embryo. Bioessays 36, 741–745 (2014).

12. Zhou, N. et al. The N-terminal domain of APJ, a CNS-based coreceptor for HIV-1, is essential for its receptor function and coreceptor activity. Virology 317, 84–94 (2003).

13. Nyimanu, D. et al. In vitro metabolism of synthetic Elabela/Toddler (ELA-32) peptide in human plasma and kidney homogenates analyzed with mass spectrometry and validation of endogenous peptide quantification in tissues by ELISA. Peptides 145, 170642 (2021).

14. Maguire, J. J., Kleinz, M. J., Pitkin, S. L. & Davenport, A. P. [Pyr1]apelin-13 identified as the predominant apelin isoform in the human heart: vasoactive mechanisms and inotropic action in disease. Hypertension 54, 598–604 (2009).

15. Yang, P. et al. Elabela/Toddler Is an Endogenous Agonist of the Apelin APJ Receptor in the Adult Cardiovascular System, and Exogenous Administration of the Peptide Compensates for the Downregulation of Its Expression in Pulmonary Arterial Hypertension. Circulation 135, 1160–1173 (2017).

16. O’Carroll, A. M., Selby, T. L., Palkovits, M. & Lolait, S. J. Distribution of mRNA encoding B78/apj, the rat homologue of the human APJ receptor, and its endogenous ligand apelin in brain and peripheral tissues. Biochim Biophys Acta 1492, 72–80 (2000).

17. Deng, C., Chen, H., Yang, N., Feng, Y. & Hsueh, A. J. W. Apela Regulates Fluid Homeostasis by Binding to the APJ Receptor to Activate Gi Signaling. J Biol Chem 290, 18261–18268 (2015).

18. Małyszko, J. et al. Cardiovascular disease and kidney transplantation–evaluation of potential transplant recipient. Polish Archives of Internal Medicine 124, (2014).

19. Pascual, M., Theruvath, T., Kawai, T., Tolkoff-Rubin, N. & Cosimi, A. B. Strategies to Improve Long-Term Outcomes after Renal Transplantation. New England Journal of Medicine 346, 580–590 (2002).

20. Woziwodzka, K. et al. Transgelin-2 in Multiple Myeloma: A New Marker of Renal Impairment? Molecules 27, 79 (2021).

21. Panaitescu, B. et al. ELABELA Plasma Concentrations are Increased in Women with Late-Onset Preeclampsia. J Matern Fetal Neonatal Med 33, 5–15 (2020).

22. Pattaro, C. et al. Estimating the glomerular filtration rate in the general population using different equations: effects on classification and association. Nephron Clin Pract 123, 102–111 (2013).

23. Hariharan, S. et al. Improved graft survival after renal transplantation in the United States, 1988 to 1996. N Engl J Med 342, 605–612 (2000).

24. First, M. R. Renal function as a predictor of long-term graft survival in renal transplant patients. Nephrology Dialysis Transplantation 18, i3–i6 (2003).

25. Kleinz, M. J. & Davenport, A. P. Immunocytochemical localization of the endogenous vasoactive peptide apelin to human vascular and endocardial endothelial cells. Regul Pept 118, 119–125 (2004).

26. Murza, A. et al. Discovery and Structure-Activity Relationship of a Bioactive Fragment of ELABELA that Modulates Vascular and Cardiac Functions. J Med Chem 59, 2962–2972 (2016).

27. Perjés, Á. et al. Characterization of apela, a novel endogenous ligand of apelin receptor, in the adult heart. Basic Res Cardiol 111, 2 (2016).

28. Xu, C. et al. ELABELA antagonizes intrarenal renin-angiotensin system to lower blood pressure and protects against renal injury. Am J Physiol Renal Physiol 318, F1122–F1135 (2020).

29. Hus-Citharel, A. et al. Effect of apelin on glomerular hemodynamic function in the rat kidney. Kidney Int 74, 486–494 (2008).

30. Gholampour, F., Bagheri, A., Barati, A., Masoudi, R. & Owji, S. M. Remote Ischemic Perconditioning Modulates Apelin Expression After Renal Ischemia-Reperfusion Injury. J Surg Res 247, 429–437 (2020).

31. Chen, H. et al. ELABELA and an ELABELA Fragment Protect against AKI. J Am Soc Nephrol 28, 2694– 2707 (2017).

32. Siddiquee, K. et al. Apelin protects against angiotensin II-induced cardiovascular fibrosis and decreases plasminogen activator inhibitor type-1 production. J Hypertens 29, 724–731 (2011).

33. Chun, H. J. et al. Apelin signaling antagonizes Ang II effects in mouse models of atherosclerosis. J Clin Invest 118, 3343–3354 (2008).

34. Chen, H. et al. Apelin protects against acute renal injury by inhibiting TGF-β1. Biochim Biophys Acta 1852, 1278–1287 (2015).

35. Sainsily, X. et al. Elabela Protects Spontaneously Hypertensive Rats From Hypertension and Cardiorenal Dysfunctions Exacerbated by Dietary High-Salt Intake. Front Pharmacol 12, 709467 (2021).

36. Małyszko, J., Małyszko, J. S., Koźminski, P. & Myśliwiec, M. Apelin and Cardiac Function in Hemodialyzed Patients: Possible Relations? AJN 26, 121–126 (2006).

37. Lacquaniti, A. et al. Apelin and copeptin: two opposite biomarkers associated with kidney function decline and cyst growth in autosomal dominant polycystic kidney disease. Peptides 49, 1–8 (2013).

38. Zhang, B., Wang, W., Wang, H., Yin, J. & Zeng, X. Promoting Effects of the Adipokine, Apelin, on Diabetic Nephropathy. PLoS One 8, e60457 (2013).

39. Lu, X. et al. Serum elabela and apelin levels during different stages of chronic kidney disease. Ren Fail 42, 667–672.

40. Schreiber, C. A., Holditch, S. J., Generous, A. & Ikeda, Y. Sustained ELABELA Gene Therapy in High-salt Diet-induced Hypertensive Rats. Curr Gene Ther 16, 349–360 (2017).

41. Chen, Z. et al. ELABELA attenuates deoxycorticosterone acetate/salt-induced hypertension and renal injury by inhibition of NADPH oxidase/ROS/NLRP3 inflammasome pathway. Cell Death Dis 11, 1–15 (2020).

42. Han, X., Wang, L.-Y., Diao, Z.-L. & Liu, W.-H. Apelin: A novel inhibitor of vascular calcification in chronic kidney disease. Atherosclerosis 244, 1–8 (2016).

43. Shan, P.-F. et al. Apelin Attenuates the Osteoblastic Differentiation of Vascular Smooth Muscle Cells. PLoS One 6, e17938 (2011).

44. Lu, X. & Hu, M. C. Klotho/FGF23 Axis in Chronic Kidney Disease and Cardiovascular Disease. Kidney Dis (Basel) 3, 15–23 (2017).

45. Li, C. & Yang, C. W. The pathogenesis and treatment of chronic allograft nephropathy. Nat Rev Nephrol 5, 513–519 (2009).

46. Daviaud, D. et al. TNFalpha up-regulates apelin expression in human and mouse adipose tissue. FASEB J 20, 1528–1530 (2006).

47. Habchi, M. et al. Circulating apelin is increased in patients with type 1 or type 2 diabetes and is associated with better glycaemic control. Clin Endocrinol (Oxf) 81, 696–701 (2014).

48. Zhang, H. et al. Serum Elabela/Toddler Levels Are Associated with Albuminuria in Patients with Type 2 Diabetes. Cell Physiol Biochem 48, 1347–1354 (2018).

49. Zhang, Y. et al. Elabela protects against podocyte injury in mice with streptozocin-induced diabetes by associating with the PI3K/Akt/mTOR pathway. Peptides 114, 29–37 (2019).

50. Ashley, E., Chun, H. J. & Quertermous, T. Opposing cardiovascular roles for the angiotensin and apelin signaling pathways. J Mol Cell Cardiol 41, 778–781 (2006).

51. Kuba, K., Sato, T., Imai, Y. & Yamaguchi, T. Apelin and Elabela/Toddler; double ligands for APJ/Apelin receptor in heart development, physiology, and pathology. Peptides 111, 62–70 (2019).

52. Ashley, E. A. et al. The endogenous peptide apelin potently improves cardiac contractility and reduces cardiac loading in vivo. Cardiovasc Res 65, 73–82 (2005).

53. Charo, D. N. et al. Endogenous regulation of cardiovascular function by apelin-APJ. Am J Physiol Heart Circ Physiol 297, H1904–1913 (2009).

54. Dai, T., Ramirez-Correa, G. & Gao, W. D. Apelin increases contractility in failing cardiac muscle. European Journal of Pharmacology 553, 222–228 (2006).

55. Farkasfalvi, K. et al. Direct effects of apelin on cardiomyocyte contractility and electrophysiology. Biochem Biophys Res Commun 357, 889–895 (2007).

